# Early Covid-19 Treatment With SARS-CoV-2 Neutralizing Antibody Sotrovimab

**DOI:** 10.1101/2021.05.27.21257096

**Authors:** Anil Gupta, Yaneicy Gonzalez-Rojas, Erick Juarez, Manuel Crespo Casal, Jaynier Moya, Diego Rodrigues Falci, Elias Sarkis, Joel Solis, Hanzhe Zheng, Nicola Scott, Andrea L. Cathcart, Christy M. Hebner, Jennifer Sager, Erik Mogalian, Craig Tipple, Amanda Peppercorn, Elizabeth Alexander, Phillip S. Pang, Almena Free, Cynthia Brinson, Melissa Aldinger, Adrienne E. Shapiro, for the COMET-ICE Investigators

**Affiliations:** Albion Finch Medical, William Osler Health Centre, Toronto, Ontario, Canada; Optimus U, Corp., Miami, Florida, USA; Continental Clinical Research, Towson, Maryland, USA; Álvaro Cunqueiro Hospital, Vigo, Pontevedra, Spain; Pines Care Research Center, Pembroke Pines, Florida, USA; Hospital de Clinicas de Porto Alegre, Porto Alegre, Brazil; Sarkis Clinical Trials, Gainesville, Florida, USA; Centex Studies, McAllen, Texas, USA; Vir Biotechnology, Inc., San Francisco, California, USA; GlaxoSmithKline, Stevenage, United Kingdom; GlaxoSmithKline, Cambridge, Massachusetts, USA; Pinnacle Research Group, Anniston, Alabama, USA; Central Texas Clinical Research, Austin, Texas, USA; Departments of Global Health and Medicine, University of Washington and Fred Hutchinson Cancer Research Center, Seattle, Washington, USA

## Abstract

**Background:** Coronavirus disease 2019 (Covid-19) disproportionately results in hospitalization and death in older patients and those with underlying comorbidities. Sotrovimab is a pan-sarbecovirus monoclonal antibody designed to treat such high-risk patients early in the course of disease, thereby preventing Covid-19 progression.

**Methods:** In this ongoing, multicenter, double-blind, phase 3 trial, nonhospitalized patients with symptomatic Covid-19 and at least one risk factor for disease progression were randomized (1:1) to an intravenous infusion of sotrovimab 500 mg or placebo. The primary efficacy endpoint was the proportion of patients with Covid-19 progression, defined as hospitalization longer than 24 hours or death, through day 29.

**Results:** In this preplanned interim analysis, which included an intent-to-treat population of 583 patients (sotrovimab, 291; placebo, 292), the primary efficacy endpoint was met. The risk of Covid-19 progression was significantly reduced by 85% (97.24% confidence interval, 44% to 96%; P = 0.002) with a total of three (1%) patients progressing to the primary endpoint in the sotrovimab group versus 21 (7%) patients in the placebo group. All five patients admitted to intensive care, including one who died by day 29, received placebo. Safety was assessed in 868 patients (sotrovimab, 430; placebo, 438). Adverse events were reported by 17% and 19% of patients receiving sotrovimab and placebo, respectively; serious adverse events were less common with sotrovimab (2%) versus placebo (6%).

**Conclusion:** Sotrovimab reduced progression of Covid-19 in patients with mild/moderate disease, was well tolerated, and no safety signals were identified.

Funded by Vir Biotechnology, Inc. and GlaxoSmithKline; ClinicalTrials.gov <u>NCT04545060</u>

## Introduction

The coronavirus disease 2019 (Covid-19) global pandemic is estimated to have killed over 3 million individuals worldwide.^1^ In the United States alone, there were an estimated 960,000 to 2.4 million hospitalizations through Fall 2020 and, at the peak of the pandemic in January 2021, 79% of intensive care hospital beds were occupied by Covid-19 patients.^1–3^ Older patients and those with certain comorbidities, such as obesity, diabetes mellitus, chronic pulmonary diseases, and chronic kidney disease, have been identified as those at highest risk of hospitalization or death.^4–8^

Highly effective therapeutics directed against severe acute respiratory syndrome coronavirus 2 (SARS-CoV-2), the virus that causes Covid-19, are urgently needed for these high-risk individuals. Recent data suggest that one option is monoclonal antibody therapy, which can reduce the risk of hospitalization^9,10^; however, the emergence and proliferation of SARS-CoV-2 variants conferring resistance to some antibodies is deeply concerning.^11,12^ Furthermore, as additional variants of concern will likely continue to emerge, there is a high unmet medical need for therapeutics that, alone or in combination, can remain effective as the virus evolves. A monoclonal antibody that neutralizes SARS-CoV-2 by targeting an evolutionarily conserved epitope that lies outside the rapidly evolving receptor-binding motif is one possible solution—it would have an intrinsically higher barrier to resistance and, due to its nonoverlapping resistance profile, could be combined with receptor-binding motif–targeted antibodies when necessary to further heighten the barrier to resistance.

Sotrovimab, formerly known as VIR-7831, is an engineered human monoclonal antibody that neutralizes SARS-CoV-2 and multiple other sarbecoviruses, including SARS-CoV, the virus responsible for the SARS outbreak two decades ago.^13^ In fact, the parental form of sotrovimab, S309, was isolated from a SARS survivor.^13^ We hypothesized that a monoclonal antibody that neutralizes all sarbecoviruses would target a highly conserved epitope that would be functionally retained as SARS-CoV-2 evolves (Fig. 1). Consistent with this hypothesis, we subsequently demonstrated that, in vitro, sotrovimab retains activity against now widespread SARS-CoV-2 variants that were first identified in the United Kingdom (B.1.1.7), South Africa (B.1.351), Brazil (P.1), and California (B.1.427/B.1.429).^11,14,15^ In contrast, many of the other monoclonal antibodies being developed for Covid-19 bind to the receptor-binding motif that engages the angiotensin-converting enzyme 2 (ACE2) receptor and is one of the most mutable and immunogenic regions of the virus; in some cases, these antibodies do not retain activity against the variants.^16–19^

**Figure 1.**
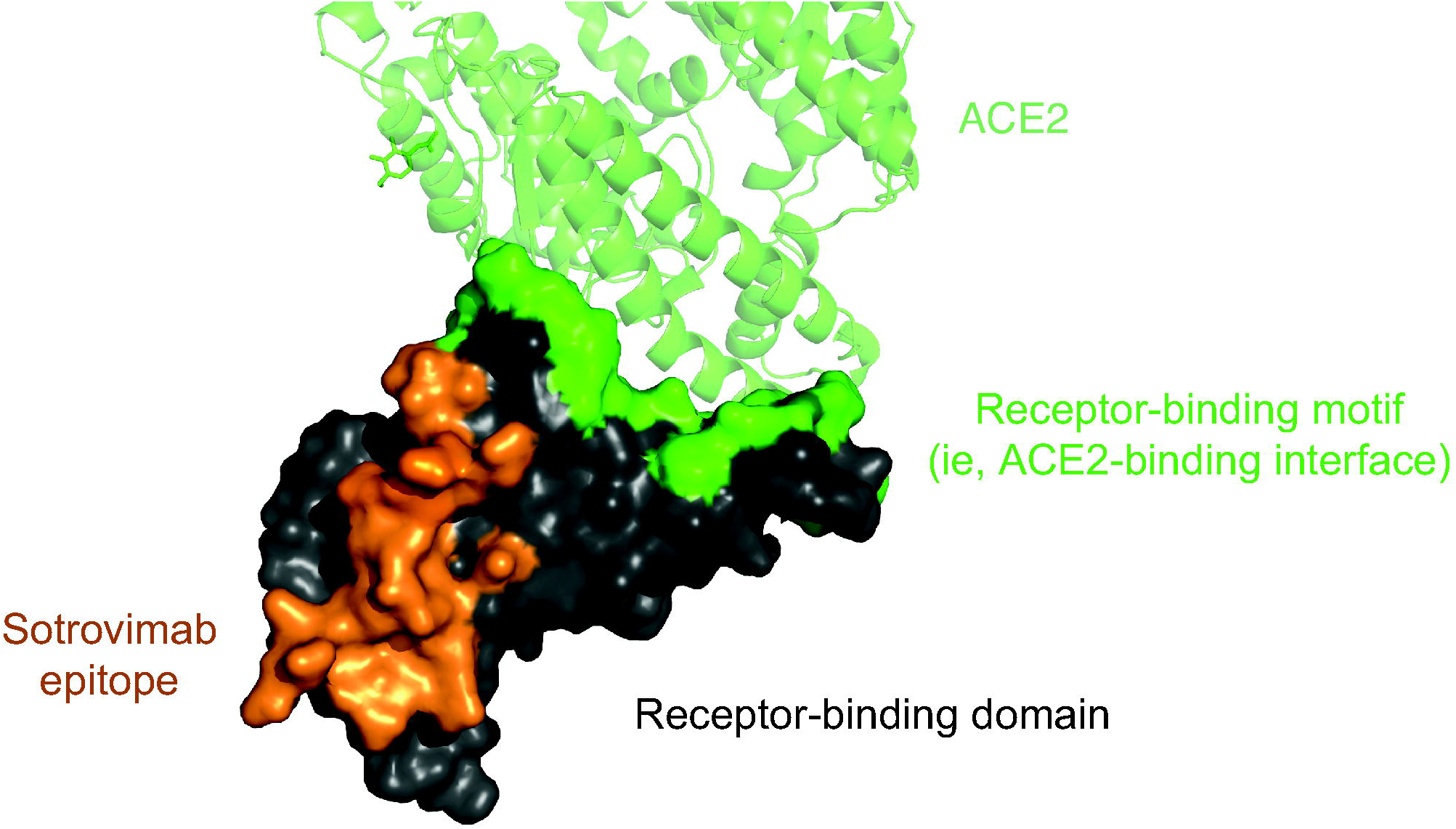
The conserved, pan-sarbecovirus binding site of sotrovimab on the spike protein of SARS-CoV-2. The SARS-CoV-2 receptor-binding domain is shown, with the ACE2 receptor-binding motif in green and the sotrovimab epitope in orange. ACE2 denotes angiotensin-converting enzyme 2.

Sotrovimab contains a two–amino acid Fc modification (termed LS) to increase half-life and potentially improve bioavailability in the respiratory mucosa through enhanced engagement with the neonatal Fc receptor.^20–22^ This modification potentially allows therapeutic concentrations for longer durations.^20–22^ Sotrovimab has demonstrated potent effector functions in vitro, which have the potential to result in immune-mediated viral clearance.^13,14^

Here, we report the results from a preplanned interim analysis of the <u>CO</u>vid-19 <u>M</u>onoclonal antibody <u>E</u>fficacy <u>T</u>rial-Intent to <u>C</u>are <u>E</u>arly (COMET-ICE) study evaluating the efficacy and safety of treatment with sotrovimab in high-risk, ambulatory patients with mild/moderate Covid-19. The trial is currently closed for enrollment; data collection is ongoing. Additional efficacy, safety, and laboratory data, as well as initial immunogenicity data, will be reported subsequently.

## Methods

### Trial Objectives and Oversight

This phase 3, randomized, double-blind, multicenter, placebo-controlled study is evaluating a single intravenous infusion of sotrovimab 500 mg for the prevention of progression of mild/moderate Covid-19 in high-risk, nonhospitalized patients. For this preplanned interim analysis, patients were recruited beginning on August 27, 2020 and followed through March 4, 2021 at 37 study sites in four countries (United States, Canada, Brazil, and Spain). Changes made to the protocol and statistical analysis plan after the trial started are summarized in the Supplementary Appendix.

This study is sponsored by Vir Biotechnology, Inc., in collaboration with GlaxoSmithKline. It is being conducted in accordance with the principles of the Declaration of Helsinki and Council for International Organizations of Medical Sciences International Ethical Guidelines, applicable International Council for Harmonisation Good Clinical Practice guidelines, and applicable laws and regulations. All patients provided written informed consent. The Sponsors designed the trial, and the Sponsors and trial investigators participated in data collection, analysis, and interpretation.

### Patients and Procedures

Adult patients 18 years of age or older with a positive reverse-transcriptase–polymerase-chain-reaction or antigen SARS-CoV-2 test result and onset of symptoms within the prior 5 days were screened for eligibility; screening was performed within 24 hours before study drug administration. Patients were required to be at high risk for Covid-19 progression, defined as older adults (age ≥55 years) or adults with at least one of the following risk factors: diabetes requiring medication, obesity (body-mass index >30 kg/m^2^), chronic kidney disease (estimated glomerular filtration rate <60 mL/min/1.73 m^2^),^23^ congestive heart failure (New York Heart Association class II or higher), chronic obstructive pulmonary disease, and moderate to severe asthma.^24^ Patients with already severe Covid-19, defined by shortness of breath at rest, respiratory distress, or requiring supplemental oxygen, were excluded. Full inclusion and exclusion criteria are described in the Supplementary Appendix.

Eligible patients were randomized 1:1 using an interactive web response system to receive either a single 500-mg, 1-hour infusion of sotrovimab or equal volume saline placebo on day 1 (Fig. 2).

**Figure 2.**
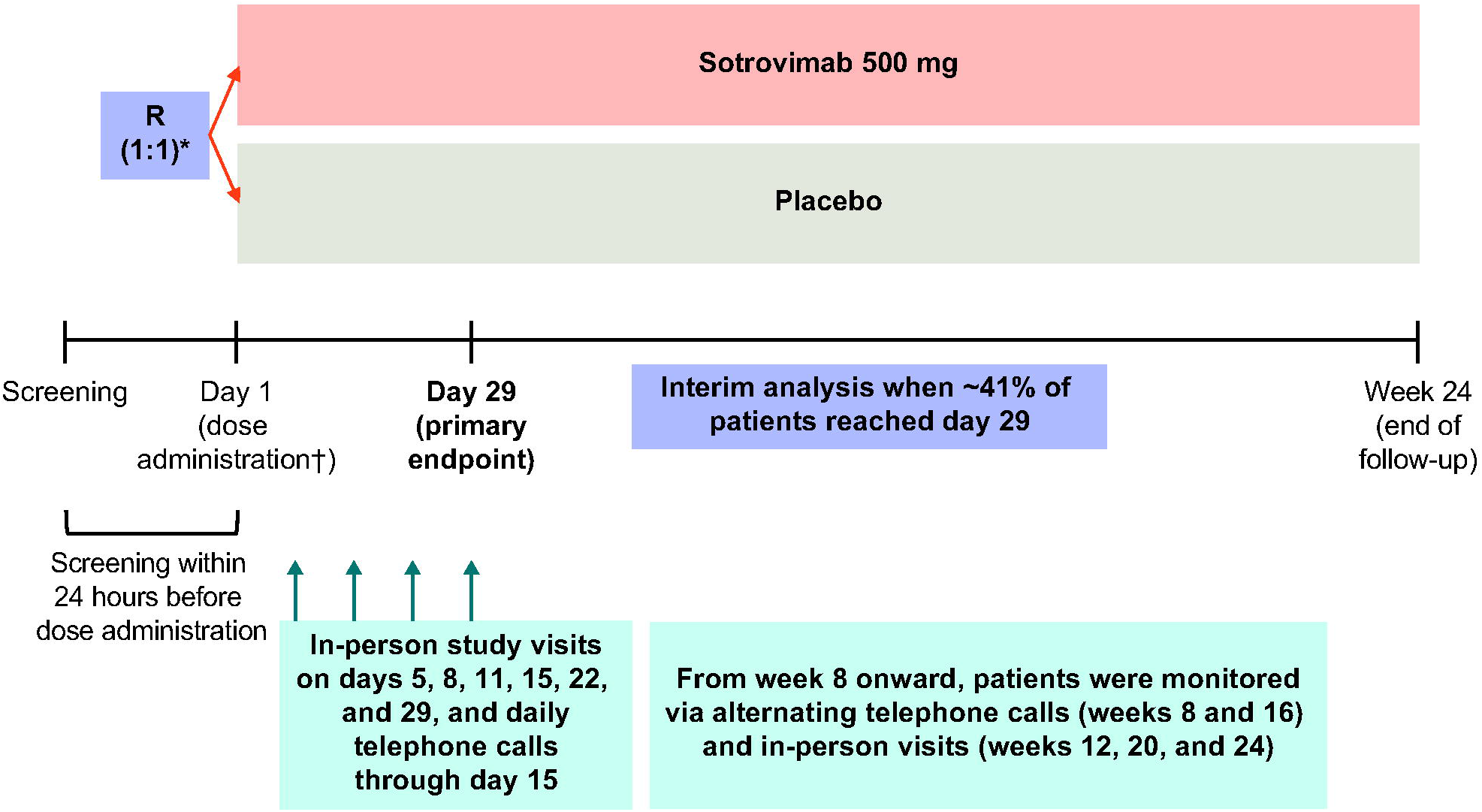
Study design. R denotes randomization. *Patients were stratified by age (≤70 vs. >70 years), symptom duration (≤3 days vs. 4-5 days), and region. †Study pharmacists reconstituted and dispensed all study medications within equal time frames to maintain blinding.

### Efficacy Assessments

The primary endpoint was the proportion of patients with hospitalization for more than 24 hours or death, due to any cause, through day 29. Secondary efficacy endpoints included the proportion of patients with an emergency room visit, hospitalization, or death; mortality; patient-reported outcomes; changes in viral load; and the proportion of patients who progressed to require supplemental oxygen.

### Safety Assessments

Safety endpoints included adverse events, serious adverse events, and adverse events of special interest, defined as infusion-related reactions (including hypersensitivity reactions), immunogenicity testing for anti-drug antibodies, and evaluation of antibody-dependent enhancement. All hospitalizations, including those due to Covid-19, were counted as serious adverse events.

### Statistical Analyses

A preplanned interim analysis for safety, futility, and profound efficacy was triggered when approximately 41% of the required number of study patients reached day 29. Sample size calculations were based on a group sequential design with two interim analyses to assess both futility due to lack of efficacy or profound efficacy. A Lan-DeMets alpha spending function to control type I error was used, employing a Pocock analog rule for futility and a Hwang-Shih-DeCani (with parameter γ = 1) analog for efficacy.^25^ The overall sample size of 1360 would have provided approximately 90% power to detect a 37.5% relative efficacy in reducing progression of Covid-19 through day 29 at the overall two-sided 5% significance level, with an assumed progression rate of 16% in the placebo group.

The interim analysis intent-to-treat (ITT) population included all randomized patients through the prespecified interim analysis cutoff date of January 19, 2021, irrespective of whether they received study drug. The interim analysis safety analysis population included all patients who received study medication and were randomized through February 17, 2021; patients were grouped according to the actual treatment received. The primary endpoint was analyzed in the ITT population using a Poisson regression model with robust sandwich estimators adjusting for treatment, duration of symptoms, age, and gender. Missing progression status was imputed under a missing at random assumption, using multiple imputation. Based on this analysis model, the statistical significance testing, the relative risk of progression, and its appropriate confidence interval (CI) are provided using the adjusted significance level for this interim analysis.

As a result of profound efficacy, an independent data monitoring committee recommended that enrollment in the study be stopped on March 10, 2021, at which time 1057 patients had been randomized. Analyses of all secondary and exploratory endpoints will be conducted when all patients have completed day 29.

## Results

### Patients

Of 795 patients screened, 583 were randomized through January 19, 2021 to sotrovimab (291 patients) or placebo (292 patients); these patients comprise the interim analysis ITT population (Fig. S1 in the Supplementary Appendix). In this ITT population, similar disposition was observed across treatment groups. Overall, four patients each in the sotrovimab and placebo groups withdrew from the study. Three patients in the sotrovimab group withdrew prior to dosing, with a fourth patient withdrawing consent on day 5 for personal reasons. One patient in the placebo group withdrew consent prior to dosing, and three patients withdrew after dosing for personal reasons (days 16, 25, and 85). The median duration of follow-up in the ITT population was 72 days (range, 5 to 190) for the sotrovimab group and 72 days (range, 16 to 190) for the placebo group.

Overall, 868 patients (sotrovimab, 430 patients; placebo, 438 patients) were randomized and received study drug through February 17, 2021; these patients comprise the interim analysis safety analysis population. The median duration of follow-up in this population was 56 days (range, 5 to 190) for the sotrovimab group and 55 days (range, 2 to 190) for the placebo group.

Treatment groups in the ITT population were well balanced for baseline demographic and disease characteristics (Table 1). Overall, 22% of patients were greater than 65 years of age, 7% were Black or African American, 63% were Hispanic or Latino, and 42% had two or more conditions considered to be risk factors for Covid-19 progression. The most common risk factors were obesity, age 55 years or older, and diabetes requiring medication. The most common presenting symptoms (>60% of all patients) were cough, muscle aches/myalgia, headache, and fatigue (Table S1 in the Supplementary Appendix). Baseline demographic and disease characteristics in the safety analysis population were similar across treatment groups and are reported in Table S2 in the Supplementary Appendix.

**Table 1.**
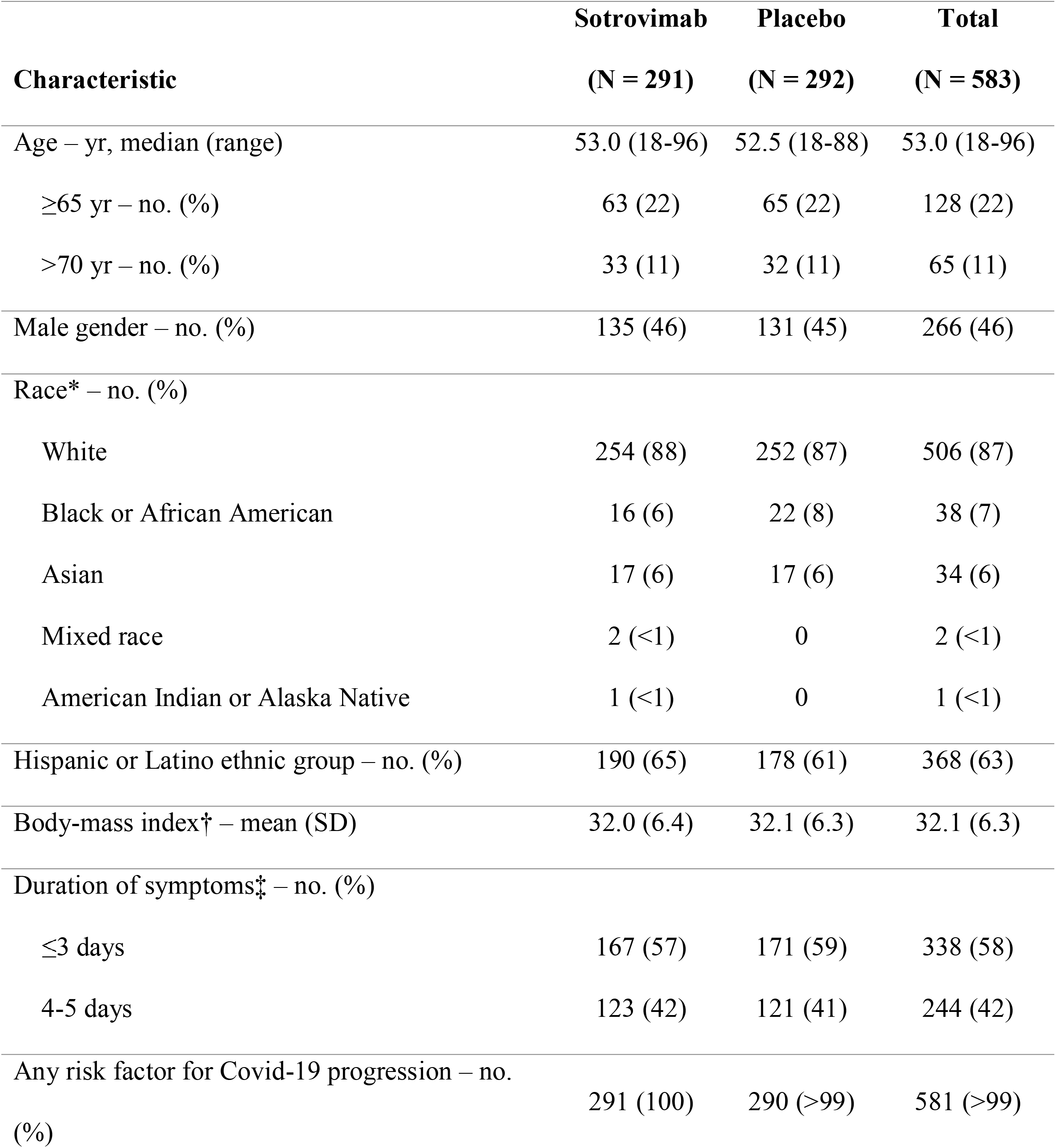
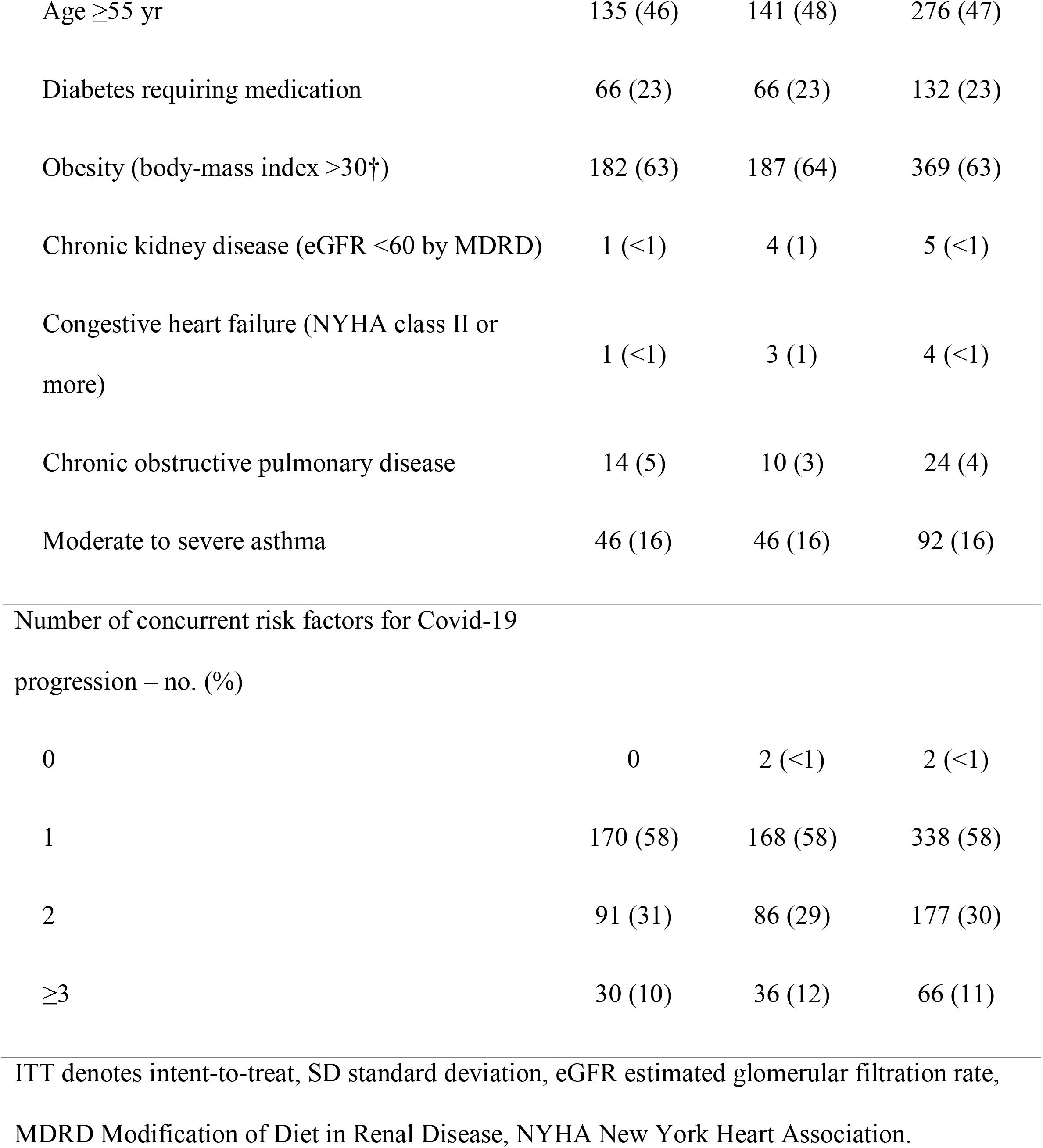
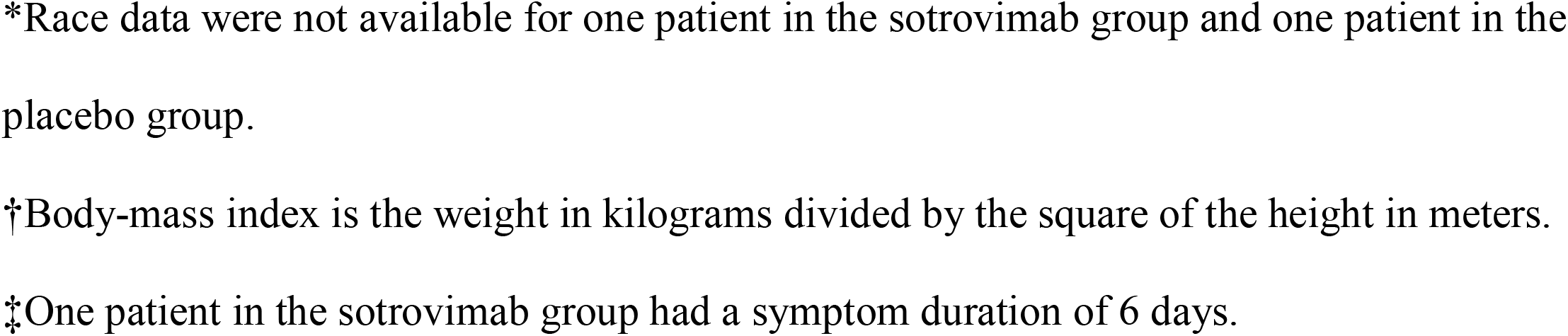
Baseline Demographic and Disease Characteristics (ITT Population)

| **Characteristic** | **Sotrovimab**<br>**(N = 291)** | **Placebo**<br>**(N = 292)** | **Total**<br>**(N = 583)** |
| --- | --- | --- | --- |
| Age – yr, median (range) | 53.0 (18-96) | 52.5 (18-88) | 53.0 (18-96) |
| ≥65 yr – no. (%) | 63 (22) | 65 (22) | 128 (22) |
| >70 yr – no. (%) | 33 (11) | 32 (11) | 65 (11) |
| Male gender – no. (%) | 135 (46) | 131 (45) | 266 (46) |
| Race* – no. (%) |  |  |  |
| White | 254 (88) | 252 (87) | 506 (87) |
| Black or African American | 16 (6) | 22 (8) | 38 (7) |
| Asian | 17 (6) | 17 (6) | 34 (6) |
| Mixed race | 2 (<1) | 0 | 2 (<1) |
| American Indian or Alaska Native | 1 (<1) | 0 | 1 (<1) |
| Hispanic or Latino ethnic group – no. (%) | 190 (65) | 178 (61) | 368 (63) |
| Body-mass index† – mean (SD) | 32.0 (6.4) | 32.1 (6.3) | 32.1 (6.3) |
| Duration of symptoms‡ – no. (%) |  |  |  |
| ≤3 days | 167 (57) | 171 (59) | 338 (58) |
| 4-5 days | 123 (42) | 121 (41) | 244 (42) |
| Any risk factor for Covid-19 progression – no.<br>(%) | 291 (100) | 290 (>99) | 581 (>99) |
| Age $\geq 55$ yr | 135 (46) | 141 (48) | 276 (47) |
| Diabetes requiring medication | 66 (23) | 66 (23) | 132 (23) |
| Obesity (body-mass index $>30^{\dagger}$) | 182 (63) | 187 (64) | 369 (63) |
| Chronic kidney disease (eGFR $<60$ by MDRD) | 1 ($<1$) | 4 (1) | 5 ($<1$) |
| Congestive heart failure (NYHA class II or more) | 1 ($<1$) | 3 (1) | 4 ($<1$) |
| Chronic obstructive pulmonary disease | 14 (5) | 10 (3) | 24 (4) |
| Moderate to severe asthma | 46 (16) | 46 (16) | 92 (16) |
| Number of concurrent risk factors for Covid-19 progression – no. (%) |  |  |  |
| 0 | 0 | 2 ($<1$) | 2 ($<1$) |
| 1 | 170 (58) | 168 (58) | 338 (58) |
| 2 | 91 (31) | 86 (29) | 177 (30) |
| $\geq 3$ | 30 (10) | 36 (12) | 66 (11) |
| ITT denotes intent-to-treat, SD standard deviation, eGFR estimated glomerular filtration rate, MDRD Modification of Diet in Renal Disease, NYHA New York Heart Association. |  |  |  |

\*Race data were not available for one patient in the sotrovimab group and one patient in the placebo group.
†Body-mass index is the weight in kilograms divided by the square of the height in meters.
‡One patient in the sotrovimab group had a symptom duration of 6 days.

### Efficacy Outcomes

Treatment with sotrovimab resulted in an 85% reduction in the need for hospitalization over 24 hours or death, due to any cause, compared with placebo (relative risk, 0.15 [97.24% CI, 0.04 to 0.56]). Three (1%) patients progressed to the primary endpoint in the sotrovimab group versus 21 (7%) patients in the placebo group (P = 0.002) (Table 2). The primary reasons for the 24 hospitalizations of more than 24 hours were consistent with progressive Covid-19 (Table S3 in the Supplementary Appendix), with one likely exception: a patient in the sotrovimab group, with a notable past medical history of small intestinal obstruction, presented 22 days after infusion with a small intestinal obstruction. Regarding the severity of these hospitalizations, all five patients who required admission to intensive care were in the placebo group; two of these five patients required invasive mechanical ventilation and a third declined intubation and subsequently died by day 29. Emergency room visits or hospitalizations for less than 24 hours were observed in fewer patients in the sotrovimab group compared with the placebo group (Table 2).

**Table 2.** Summary of Efficacy Outcomes Through Day 29 (ITT Population)

|  | Sotrovimab | Placebo |
| --- | --- | --- |
|  | (N = 291) | (N = 292) |
| Primary outcome |  |  |
| Hospitalized >24 hours or death for any cause – no. (%) | 3 (1) | 21 (7) |
| Hospitalized >24 hours for any cause | 3 (1) | 21 (7) |
| Death by any cause | 0 | 1 (<1) |
| Alive and not hospitalized – no. (%) | 284 (98) | 270 (92) |
| Missing – no. (%) | 4 (1) | 1 (<1) |
| Withdrew consent prior to dosing – no. (%) | 3 (1) | 1 (<1) |
| Percent reduction (97.24% CI), P value | 85% (44% to 96%); P = 0.002 |  |
| Other clinical outcomes* |  |  |
| Emergency room visit, hospitalization, or death for any cause – no. (%) | 6 (2) | 28 (10) |
| Emergency room visit for any cause | 2 (<1) | 8 (3) |
| Hospitalized for any cause | 4 (1)† | 21 (7) |
| Death by any cause | 0 | 1 (<1) |
| Emergency room visit without hospitalization or hospitalized for <24 hours for any cause – no. (%) | 3 (1) | 7 (2) |
| Severe or critical progression‡ – no. (%) | 2 (<1) | 19 (7) |
| Low-flow nasal cannula or face mask | 2 (<1) | 11 (4) |
| Non-rebreather mask, high-flow nasal cannula, or<br>noninvasive ventilation | 0 | 5 (2) |
| Invasive mechanical ventilation | 0 | 2 (<1) |
| Death by any cause | 0 | 1 (<1) |
| Admission to intensive care for any cause – no. (%) | 0 | 5 (2) |

ITT denotes intent-to-treat, CI confidence interval.
\*Inferential testing of secondary endpoints has not been performed at this interim analysis.
†One patient was hospitalized for less than 24 hours for diabetes management.
‡Severe or critical progression as manifested by supplemental oxygen use.

### Safety

The proportion of patients in the safety analysis population who reported an adverse event was 17% (73 of 430 patients) in the sotrovimab group and 19% (85 of 438 patients) in the placebo group (Table 3). A lower proportion of patients reported grade 3 or 4 adverse events in the sotrovimab group (2%) compared with the placebo group (6%). Overall, the only adverse event occurring in at least 1% of patients receiving sotrovimab was diarrhea, which occurred infrequently—six (1%) patients in the sotrovimab group versus three (<1%) patients in the placebo group. Among patients in the sotrovimab group, all cases of diarrhea were mild (five patients) or moderate (one patient) in severity.

**Table 3.**
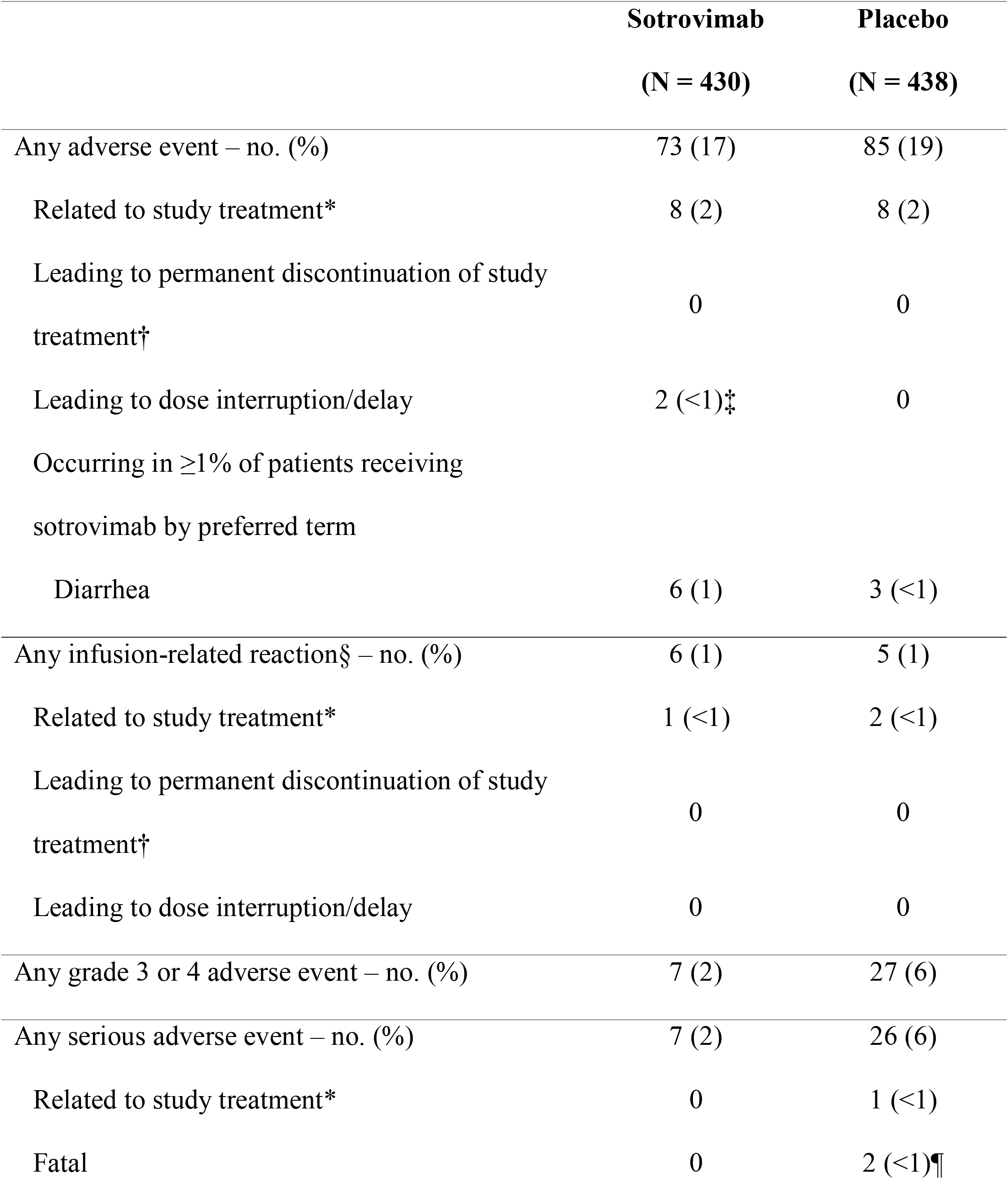
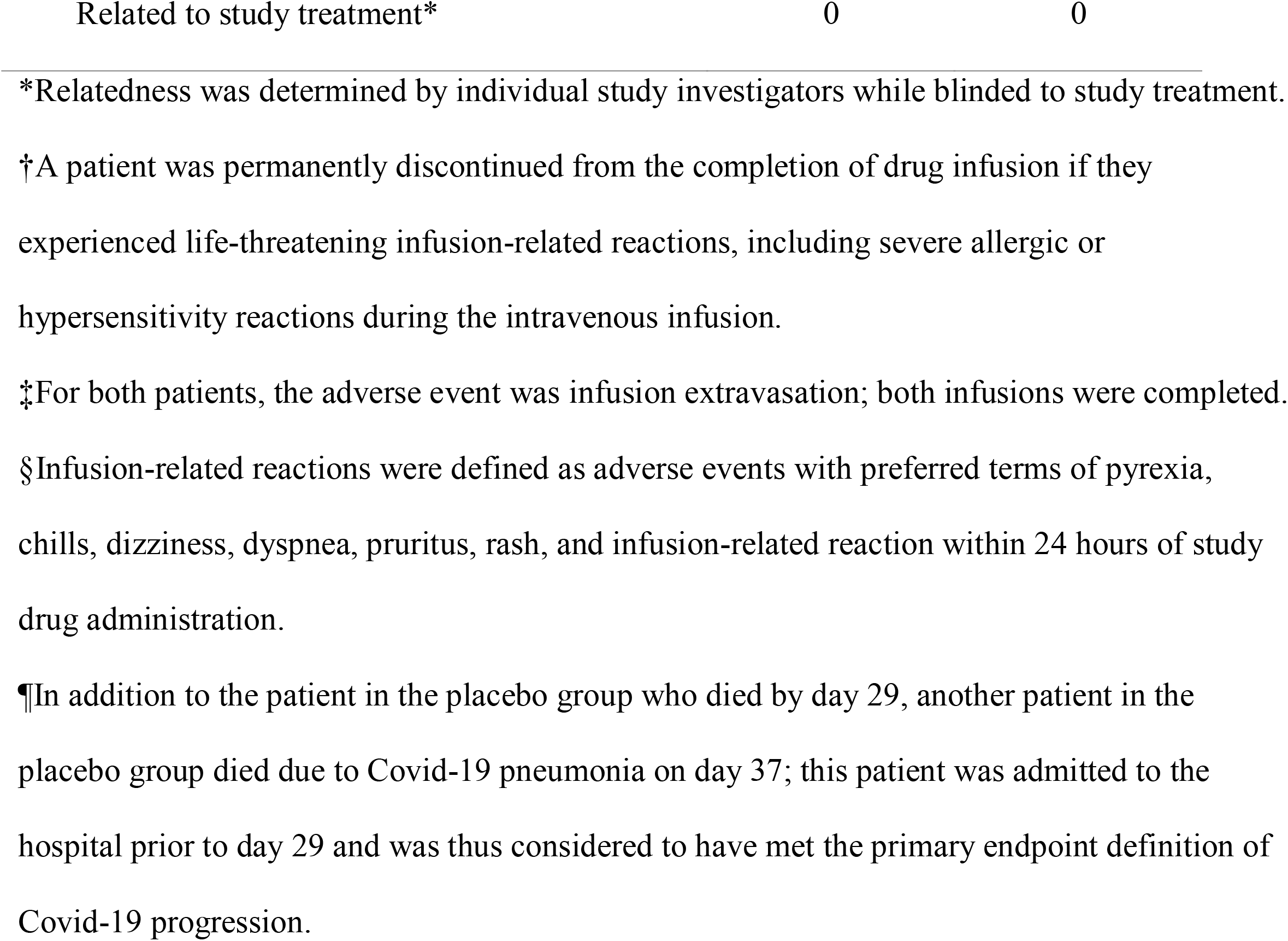
Summary of Adverse Events (Safety Analysis Population)

Infusion-related reactions were observed in a similar proportion of patients receiving sotrovimab (1%) compared with placebo (1%). One patient receiving sotrovimab had an infusion-related reaction that was considered related to study treatment: moderate (grade 2) dyspnea.

Serious adverse events occurred in 2% of patients receiving sotrovimab and 6% of patients receiving placebo. Most of these events were hospitalizations due to Covid-19–related causes. No serious adverse events were considered related to sotrovimab. One patient in the placebo group died after day 29; this patient died due to Covid-19 pneumonia on day 37.

No trends were observed in hematologic, liver, or chemistry laboratory data. Overall, laboratory results were similar in the sotrovimab and placebo groups.

## Discussion

In this preplanned interim analysis of the COMET-ICE study, a single 500-mg dose of sotrovimab profoundly reduced the risk of hospitalization (>24 hours) or death in high-risk adults with symptomatic Covid-19 by 85% compared with placebo (P = 0.002). For every 17 high-risk patients with symptomatic Covid-19, sotrovimab prevented one hospitalization. Importantly, among those who were hospitalized, no patient who received sotrovimab required admission to intensive care compared with five patients who received placebo, suggesting that sotrovimab may also prevent more severe complications of Covid-19 in addition to preventing the need for hospitalization itself. Furthermore, as a result of investigator site selection, over 60% of the study population consisted of patients self-identifying as Hispanic or Latino; thus, this trial is one of the first to demonstrate efficacy in a population that has been largely underrepresented in Covid-19 clinical trials, despite the disproportionately negative impact the pandemic has had in this ethnic group. Overall, sotrovimab was well tolerated, and no safety signals were identified in this study. There was also no evidence of antibody-dependent enhancement with sotrovimab, which would have manifested as worsening of disease compared with placebo.^26^

Sotrovimab is thus a potentially critical therapeutic in the fight against Covid-19, for which there remains a high unmet medical need despite the recent success of preventative measures, such as vaccines. Challenges with access to vaccines, vaccine hesitancy, medical contraindications to vaccines, immunocompromised individuals who may not respond to a vaccine, and most importantly, the potential emergence of variant viruses that escape vaccine-derived immunity, will all contribute to what is likely to be an unfortunately large and enduring number of Covid-19 cases in need of treatment.

Importantly, treatments for Covid-19 will need to retain activity even in the face of a rapidly evolving virus. To that end, sotrovimab was selected to have an intrinsically high barrier to resistance as a result of targeting a pan-sarbecovirus epitope.^14^ In one analysis, among more than 584,000 sequences in the Global Influenza Surveillance and Response System database (Global Initiative on Sharing All Influenza Data), amino acid positions comprising the sotrovimab epitope were at least 99.96% conserved in naturally occurring viruses.^14^ Moreover, when necessary to further enhance breadth and barrier to resistance, sotrovimab can likely be combined with currently authorized receptor-binding motif–targeted antibodies due to its nonoverlapping resistance profile.

This interim report has three major limitations. First, with only three hospitalizations in the sotrovimab group, it is not possible to determine which patient or disease characteristics might be associated with sotrovimab treatment failure. Second, the safety dataset was moderate in size (430 patients) and thus a rare (much less than 1%) adverse event may not have been observed, though one would not be expected as sotrovimab was derived from an antibody isolated from a recovered SARS-CoV patient, has minimal engineering, and targets a viral epitope (not a host epitope). Third, secondary and exploratory endpoint analyses are excluded from this interim report since the study is ongoing; such analyses are needed to further determine the potential additional benefits of sotrovimab.

This study has two key scientific and medical ramifications, beyond demonstrating the therapeutic value of sotrovimab. First, the results indicate that a non–receptor-binding motif binding antibody, which does not directly block the ACE2 receptor interaction, can be clinically therapeutic, and suggests a role for other receptors.^27^ Second, as sotrovimab has potent effector function, the absence of safety signals and profound efficacy strongly suggest that effector function is neither detrimental nor associated with antibody-dependent enhancement.^26^ In fact, preclinical models of Covid-19 suggest that its potent effector function may be beneficial.^13,14^

In conclusion, results from this interim analysis of the COMET-ICE trial strongly support that sotrovimab can be an important therapeutic for the outpatient treatment of Covid-19. Notably, a 500-mg dose may also enable intramuscular administration, increasing the convenience of and access to antibody therapeutics for patients with Covid-19. Studies are currently underway to evaluate this route of administration. Importantly, as a component of a comprehensive pandemic preparedness strategy, a pan-sarbecovirus antibody has the potential to be an off-the-shelf, ready-to-deploy therapeutic for future coronavirus pandemics.

## Supporting information

Supplemental Fil

CONSORT Checklist

## Data Availability

This is an interim analysis; the data will not be made available at this time.

## Acknowledgments

Medical writing support was provided by Caryn Gordon, PharmD, and Courtney St. Amour, PhD, of Cello Health Communications/SciFluent, and was funded by Vir Biotechnology, Inc. We thank Krystyna Grycz, Jordan Clark, and Minnie Kuo, of Vir Biotechnology, Inc., for clinical operations support.

