## Supplemental Fil for "Early Covid-19 Treatment With SARS-CoV-2 Neutralizing Antibody Sotrovimab"

**SUPPLEMENTARY APPENDIX**

**Table of Contents**

| List of investigators | Page 2 |
| --- | --- |
| Supplementary methods | Page 6 |
| Changes to protocol/SAP | Page 12 |
| Table S1. Presenting symptoms (ITT population) | Page 17 |
| Table S2. Baseline demographic and disease characteristics (safety analysis population) | Page 18 |
| Table S3. Primary reasons for hospitalizations of more than 24 hours (ITT population) | Page 21 |
| Figure S1. Patient disposition (ITT population) | Page 24 |

**List of Investigators**

Haider Afzal – Inquest Clinical Research, Baytown, TX, USA

Hassan Ali – Allied Biomedical Research Institute, Miami, FL, USA

German Alvarez – Model Research Center, LLC, Tampa, FL, USA

Duane Anderson – Excel Clinical Research, Las Vegas, NV, USA

Masoud Azizad – Valley Clinical Trials, Northridge, CA, USA

Ali Bajwa – Centex Studies - Westfield, Houston, TX, USA

Greg Bostick – Cullman Clinical Trials, Cullman, AL, USA

Cynthia Brinson – Central Texas Clinical Research, Austin, TX, USA

Jorge Caso – Angels Clinical Research Institute - International Research Consultants, Miami, FL, USA

Edward M. Cordasco – Remington-Davis, Inc., Columbus, OH, USA

Manuel Crespo Casal – Álvaro Cunqueiro Hospital, Vigo, Pontevedra, Spain

Armando Curra – Miramax Clinical Research, North Miami, FL, USA

Vinícius Buaes Dal Maso - Instituto Méderi de Pesquisa e Saúde, Passo Fundo, Rio Grande do Sul, Brazil

Carlos Augusto de Aguiar Quadros – Unidade Hospital Leforte Christóvão da Gama, Santo Andre, Sao Paulo, Brazil

Jorge Diaz – private practice, Hialeah, FL, USA

Ankur Doshi – PrimeCare Medical Group, Houston, TX, USA

Victor Escobar – 1960 Family Practice, PA, Houston, TX, USA

Ladynez Espinal – AdMed Research, Miramar, FL, USA

Diego Rodrigues Falci – Hospital de Clínicas de Porto Alegre, Porto Alegre, Rio Grande do Sul, Brazil

Adil Fatakia – Tandem Clinical Research, Marrero, LA, USA

Alfredo Fernandez – Clinical Trials of Tampa, Tampa, FL, USA

Augusto Focil – FOMAT Medical Research, Oxnard, CA, USA

Almena Free – Pinnacle Research Group, LLC, Anniston, AL, USA

Marcy Goisse – Frontier Clinical Research, LLC, Smithfield, PA, USA

Yaneicy Gonzalez-Rojas – Optimus U Corporation, Miami, FL, USA

Linda Gorgos – AXCES Research Group, LLC, Santa Fe, NM, USA

Anil Gupta – Albion Finch Medical, William Osler Health Centre, Toronto, ON, Canada

Luis Hernandez – Innovation Medical Research Center, Palmetto Bay, FL, USA

Rubaba (Ruby) Hussain – RH Medical Urgent Care, Bronx, NY, USA

Kimball Johnson - iResearch Atlanta, LLC, Decatur, GA, USA

Erick Juarez – Continental Clinical Research, Towson, MD, USA

John Kowalczyk – American Institute of Research, Los Angeles, CA, USA

Glenn Leavitt – Leavitt Clinical Research, Idaho Falls, ID, USA

Raymond Little – Houston Heart & Vascular Associates, Humble, TX, USA

Kleber Luz – Centro de Estudos e Pesquisas em Moléstias Infecciosas Ltda, Natal, Rio Grande do Norte, Brazil

Luis Martinez – Universal Axon Clinical Research, LLC, Doral, FL, USA

Shilpi Mittal – Care United Research, LLC, Forney, TX, USA

Bharat Mocherla – Las Vegas Medical Research, Las Vegas, NV, USA

Jaynier Moya – Pines Care Research Center, LLC, Pembroke Pines, FL, USA

Silvia Narejos Perez – Cap Centelles, Centelles, Spain

Thinh Nguyen – Buckhead Primary Care Research, Atlanta, GA, USA

John O’Mahony – Bluewater Clinical Research Group, Inc, Sarnia, ON, Canada

Naval Parikh – Napa Research, LLC, Pompano Beach, FL, USA

Russell Perry – Gadolin Research, LLC, Beaumont, TX, USA

Ronald Pucillo – Javara Research, Sugar Land, TX, USA

Eduardo Ramacciotti – Unidade Leforte Morumbi, Vila Assuncao, São Paulo, Brazil

Larry Reed – Healthcare Research Network, Hazelwood, MO, USA

Juan Roldan Sanchez – Institut Català de Serveis Mèdics, Girona, Spain

Peter Ruane – Ruane Clinical Research Group, Inc., Los Angeles, CA, USA

Yessica Sachdeva – The Institute for Liver Health, Mesa, AZ, USA

Elias H. Sarkis – Sarkis Clinical Trials, Gainesville, FL, USA

Michael Seep – Centex Studies, Lake Charles, LA, USA

Patricia Segura – Hospital Nacional Hipolito Unanue, El Agustino, Lima, Peru

Adrienne Shapiro – Fred Hutchinson Cancer Research Center, Seattle, WA, USA

Lawrence Sher – Peninsula Research Associates, Inc., Rolling Hills Estates, CA, USA

Joel Solis – Centex Studies, McAllen, TX, USA

Guillermo Somodevilla – Cordova Research Institute, Miami, FL, USA

Claudio Marcel Stadnik – Santa Casa de Misericórdia de Porto Alegre, Porto Alegre, Rio Grande do Sul, Brazil

Luis Zepeda – Vilo Research Group, Inc., Houston, TX, USA

**Supplementary Methods**

**Inclusion Criteria**

Patients are eligible to be included in the study only if all of the following criteria apply:

*Age and Risk Factors*

- Patient must be 18 years of age or older AND at high risk of progression of Covid-19 based on presence of one or more of the following risk factors:
  - Diabetes (requiring medication)
  - Obesity (body-mass index >35 kg/m^2^)
    - Change: body-mass index threshold was >30 kg/m^2^ in the original protocol. See “Changes to Protocol/SAP” section for more information
  - Chronic kidney disease (i.e., estimated glomerular filtration rate <60 mL/min/1.73 m^2^ according to the Modification of Diet in Renal Disease study equation)
  - Congestive heart failure (New York Heart Association class II or more)
  - Chronic obstructive pulmonary disease (history of chronic bronchitis, chronic obstructive lung disease, or emphysema with dyspnea on physical exertion) and moderate to severe asthma (patient requires an inhaled steroid to control symptoms or has been prescribed a course of oral steroids in the past year)

**or**

- Patient 55 years of age or older, irrespective of comorbidities

Note: target enrollment of ~15% of patients over 70 years of age

Change: target enrollment criterion was not in the original protocol. See “Changes to Protocol/SAP” section for more information

*Type of Patient and Disease Characteristics*

- Patients who have a positive SARS-CoV-2 test result (by any validated diagnostic test [e.g. RT-PCR, antigen-based testing on any specimen type])

**and**

- Oxygen saturation ≥94% on room air

**and**

- Have Covid-19 defined by one or more of the following symptoms: fever, chills, cough, sore throat, malaise, headache, joint or muscle pain, change in smell or taste, vomiting, diarrhea, shortness of breath on exertion

**and**

- Less than or equal to 5 days from onset of symptoms

*Sex and Contraceptive/Barrier Requirements*

- No gender restrictions
- Female patients must meet and agree to abide by the following contraceptive criteria:
  - Contraception use by women should be consistent with local regulations regarding the methods of contraception for those participating in clinical studies
  - A female patient is eligible to participate if she is not pregnant or breastfeeding and one of the following conditions applies:
- Is a woman of nonchildbearing potential
- Is a woman of childbearing potential and is using a contraceptive method that is highly effective, with a failure rate of <1%, during the study intervention period and for up to 24 weeks after the last dose of study intervention. The investigator should evaluate potential for contraceptive method failure (e.g., noncompliance, recently initiated) in relationship to the first dose of study intervention
- A woman of childbearing potential must have a negative highly sensitive pregnancy test (urine or serum as required by local regulations) before the first dose of study intervention. If a urine test cannot be confirmed as negative (e.g., an ambiguous result), a serum pregnancy test is required. In such cases, the patient must be excluded from participation if the serum pregnancy result is positive
- The investigator is responsible for the review of medical history, menstrual history, and recent sexual activity to decrease the risk for inclusion of a woman with an early undetected pregnancy

**Informed Consent**

- Capable of giving signed informed consent, which includes compliance with the requirements and restrictions listed in the informed consent form and in this protocol

**or**

- If patients are not capable of giving written informed consent, alternative consent procedures will be followed

**Exclusion Criteria**

- Patients are excluded from the study if any of the following criteria apply:

*Medical Conditions*

- Currently hospitalized or judged by the investigator as likely to require hospitalization in the next 24 hours
- Symptoms consistent with severe Covid-19 as defined by shortness of breath at rest or respiratory distress or requiring supplemental oxygen
- Patients who, in the judgment of the investigator, are likely to die in the next 7 days
- Severely immunocompromised patients, including but not limited to cancer patients actively receiving immunosuppressive chemotherapy or immunotherapy, those with a solid organ transplant or allogeneic stem cell transplant within the last 3 months, or those having conditions requiring the use of systemic corticosteroids equivalent to ≥0.5 mg/kg of body weight per day of prednisone within 6 weeks of randomization
  - Change: this criterion has been modified from the original protocol to clarify the definition of “severely immunocompromised.” See the “Changes to Protocol/SAP” section for more information
- Known hypersensitivity to any constituent present in the investigational product
- Previous anaphylaxis or hypersensitivity to a monoclonal antibody

*Prior/Concurrent Clinical Study Experience*

- Enrollment in any investigational vaccine study within the last 180 days or any other investigational drug study within 30 days prior to day 1 or within five half-lives of the investigational compound, whichever is longer
- Enrollment in any trial of an investigational vaccine for SARS-CoV-2

*Other Exclusions*

- Receipt of any vaccine within 48 hours prior to enrollment. Receipt of a SARS-CoV-2 vaccine prior to randomization at any time point. Vaccination (including vaccination for SARS-CoV-2) will not be allowed for 4 weeks after dosing
  - Change: modified this criterion to clarify restrictions around dosing with a SARS-CoV-2 vaccine. See the “Changes to Protocol/SAP” section for more information
- Receipt of convalescent plasma from a recovered Covid-19 patient or anti–SARS-CoV-2 monoclonal antibody within the last 3 months
- Patients who, in the judgment of the investigator, will be unlikely or unable to comply with the requirements of the protocol through day 29

**Procedures**

In-person study visits occurred on days 5, 8, 11, 15, 22, and 29, as well as daily telephone calls through day 15 (except on in-person visit days) to assess for adverse events and worsening of Covid-19. Starting at week 8, patients were monitored monthly via alternating telephone calls (weeks 8 and 16) and in-person visits (weeks 12, 20, and 24) for Covid-19 illness for a total of 24 weeks from dosing. Blood samples were collected for laboratory assessments at days 1, 15, 22, and 29 and for anti-drug antibodies on days 1 and 29. Patient-reported outcome assessments are being administered through week 24.

**Changes to Protocol/SAP**

- Local safety laboratory assessments at screening were modified to specify that only ABO typing was required per protocol
- Rationale:
- While certain screening assessments to confirm study patient eligibility must be performed or documented locally (e.g., pregnancy test for women of childbearing potential and SARS-CoV-2 infection via a validated diagnostic test), ABO typing is the only required local laboratory assessment at screening for safety purposes
- All other local safety laboratory assessments during the screening visit (hematology, clinical chemistry, and coagulation parameters) may or may not be performed according to the clinical discretion of the study investigator or as required by local regulations
- Considering the time to randomization and requirement for central laboratory testing on day 1 (randomization), the Sponsor felt that the amount of sample collection was overly cumbersome for sites
- Study objectives and endpoints (secondary and exploratory endpoints) were modified, including the introduction of an additional secondary endpoint (i.e., evaluation of the proportion of patients who have progression of Covid-19 through day 29 as defined by a visit to a hospital emergency room for management of illness, hospitalization for acute management of illness, or death), amendment to existing patient-reported outcome and virology endpoints, and introduction of additional exploratory endpoints
  - Rationale:
  - Preliminary clinical trial data generated for other monoclonal antibodies targeting Covid-19 suggest that the rate of progression of Covid-19 leading to the requirement for hospitalization may be lower than originally estimated and the efficacy of antispike protein monoclonal antibodies in the treatment of Covid-19 in the outpatient population is greater than a 50% decrease in the proportion of medically attended visits through day 29
  - In light of new data from recent clinical trials of anti–SARS-CoV-2 mAbs, the Sponsor has modified the secondary and exploratory endpoints to reflect the clinical outcomes that are most relevant in the outpatient populations who receive Covid-19 monoclonal antibodies
- Modified the “Age and Risk Factors” eligibility criteria (inclusion criteria #3 and #4) to enrich high-risk populations with the highest unmet medical need
  - Rationale:
  - The “Age and Risk Factors” eligibility criteria has been modified to increase the body-mass index requirement defining obesity and introduce a targeted minimum number of patients (approximately 15%) >70 years of age
  - In light of emerging clinical data on the progression of Covid-19 in the outpatient setting, the Sponsor has amended key eligibility criteria to enrich for populations at high risk for progression to severe Covid-19
  - Modified the “Medical Conditions” eligibility criteria (exclusion criteria #10) to enrich high-risk populations with the highest unmet medical need
  - Rationale:
  - The “Medical Conditions” eligibility criteria have been modified to clarify the definition of “severely immunocompromised”
  - In light of emerging clinical data on the progression of Covid-19 in the outpatient setting, the Sponsor has modified key eligibility criteria to enrich for immunosuppressed populations at high risk for progression to severe Covid-19
- Modified the “Other Exclusions” eligibility criteria and “Medication Not Permitted During the Study” to clarify restrictions around dosing with a SARS-CoV-2 vaccine
  - Rationale:
    - Considering the recent data published on SARS-CoV-2 vaccines, the Sponsor has amended the protocol to clarify restrictions surrounding administration of an experimental or approved SARS-CoV-2 vaccine
    - Clarified guidance for infusion-related reactions
      - Rationale:
        - Protocol language was amended to clarify that sites should follow local or institutional guidelines for the treatment of infusion-related reactions
        - Statistical analysis plan (interim analysis #1, interim analysis #2, and statistical approach) were modified

Rationale:

Preliminary clinical trial data generated for other monoclonal antibodies targeting Covid-19 suggest that the rate of progression of Covid-19 to requirement for hospitalization may be lower than originally estimated and the efficacy of antispike protein monoclonal antibodies in the treatment of Covid-19 in the outpatient population is greater than a 50% decrease in the proportion of medically attended visits through day 29

In light of this emerging data, VIR Biotechnology, Inc./GlaxoSmithKline has modified the interim analyses to more accurately reflect the most recent data on progression of Covid-19 and the potential efficacy of monoclonal antibodies against SARS-CoV-2

Specified an intention to conduct a “non–Covid-19” safety analysis

Rationale:

As noted in Protocol Section 8.4, since it will not be possible to delineate in a single patient whether the hospitalization is directly related to Covid-19 complications or could be related to sotrovimab causing more severe disease due to antibody-dependent enhancement, all hospitalizations regardless of cause will be included in the primary endpoint and will also be counted as serious adverse events

To inform on the number and nature of non–Covid-19 adverse events and serious adverse events, additional safety analyses will be performed in which select, prespecified terms consistent with known progression of Covid-19 disease will be excluded. Details of these and all analyses, including example outputs, will be documented in the analysis plan

**Table S1. Presenting Symptoms (ITT Population)**

| Symptom – no. (%) | Sotrovimab  (N = 291) | Placebo  (N = 292) | Total  (N = 538) |
| --- | --- | --- | --- |
| Cough | 240 (82) | 247 (85) | 487 (84) |
| Muscle aches/myalgia | 215 (74) | 215 (74) | 430 (74) |
| Headache | 202 (69) | 216 (74) | 418 (72) |
| Fatigue | 180 (62) | 183 (63) | 363 (62) |
| Malaise | 172 (59) | 172 (59) | 344 (59) |
| Sore throat | 171 (59) | 172 (59) | 343 (59) |
| Fever | 164 (56) | 168 (58) | 332 (57) |
| Loss of taste | 171 (59) | 159 (54) | 330 (57) |
| Loss of smell | 175 (60) | 152 (52) | 327 (56) |
| Chills | 164 (56) | 158 (54) | 322 (55) |
| Joint pain/arthralgia | 153 (53) | 153 (52) | 306 (52) |
| Shortness of breath | 131 (45) | 131 (45) | 262 (45) |
| Diarrhea | 87 (30) | 101 (35) | 188 (32) |
| Nausea | 85 (29) | 91 (31) | 176 (30) |
| Vomiting | 34 (12) | 37 (13) | 71 (12) |

ITT denotes intent-to-treat.

**Table S2. Baseline Demographic and Disease Characteristics (Safety Analysis Population)**

| Characteristic | Sotrovimab  (N = 430) | Placebo  (N = 438) | Total  (N = 868) |
| --- | --- | --- | --- |
| Age – yr, median (range) | 53.0 (18-96) | 52.0 (17-88) | 53.0 (17-96) |
| ≥65 yr – no. (%) | 84 (20) | 88 (20) | 172 (20) |
| >70 yr – no. (%) | 42 (10) | 42 (10) | 84 (10) |
| Male gender – no. (%) | 194 (45) | 212 (48) | 406 (47) |
| Race* – no. (%) |  |  |  |
| White | 374 (87) | 384 (88) | 758 (88) |
| Black or African American | 27 (6) | 33 (8) | 60 (7) |
| Asian | 21 (5) | 19 (4) | 40 (5) |
| Mixed race | 6 (1) | 0 | 6 (<1) |
| American Indian or Alaska Native | 1 (<1) | 1 (<1) | 2 (<1) |
| Hispanic or Latino ethnic group – no. (%) | 280 (65) | 280 (64) | 560 (65) |
| Body-mass index† – mean (SD) | 32.1 (6.4) | 32.5 (6.7) | 32.3 (6.5) |
| Duration of symptoms‡ – no. (%) |  |  |  |
| ≤3 days | 254 (59) | 260 (59) | 514 (59) |
| 4-5 days | 173 (40) | 178 (41) | 351 (40) |
| Any risk factor for Covid-19 progression – no. (%) | 427 (>99) | 434 (>99) | 861 (>99) |
| Age ≥55 yr | 195 (45) | 205 (47) | 400 (46) |
| Diabetes requiring medication | 93 (22) | 88 (20) | 181 (21) |
| Obesity (body-mass index >30†) | 267 (62) | 292 (67) | 559 (64) |
| Chronic kidney disease (eGFR <60 by MDRD) | 2 (<1) | 5 (1) | 7 (<1) |
| Congestive heart failure (NYHA class II or more) | 4 (<1) | 3 (<1) | 7 (<1) |
| Chronic obstructive pulmonary disease | 24 (6) | 18 (4) | 42 (5) |
| Moderate to severe asthma | 69 (16) | 72 (16) | 141 (16) |
| Number of concurrent risk factors for Covid-19 progression – no. (%) |  |  |  |
| 0 | 3 (<1) | 4 (<1) | 7 (<1) |
| 1 | 251 (58) | 250 (57) | 501 (58) |
| 2 | 132 (31) | 130 (30) | 262 (30) |
| ≥3 | 44 (10) | 54 (13) | 98 (11) |

SD denotes standard deviation, eGFR estimated glomerular filtration rate, MDRD Modification of Diet in Renal Disease, NYHA New York Heart Association.
*Race data were not available for one patient in the sotrovimab group and one patient in the placebo group.

†Body-mass index is the weight in kilograms divided by the square of the height in meters.

‡One patient in the sotrovimab group had a symptom duration of 6 days. For two other patients in the sotrovimab group, symptom duration data were not available at the time of this interim analysis.

**Table S3. Primary Reasons for Hospitalizations of More Than 24 Hours (ITT Population)**

| Patient | Age range (yr) | Hospitalization day | Primary reason | Intensive care unit admission | Invasive mechanical ventilation | Fatal |
| --- | --- | --- | --- | --- | --- | --- |
| Sotrovimab-treated patients | | | | | | |
| A | >95 to ≤100 | 19 | Covid-19 | N | N | N |
| B | >60 to ≤65 | 22 | Small intestinal obstruction | N | N | N |
| C | >70 to ≤75 | 2 | Covid-pneumonia | N | N | N |
| Placebo-treated patients | | | | | | |
| D | >50 to ≤55 | 4 | Covid-pneumonia | N | N | N |
| E | >45 to ≤50 | 4* | Covid-pneumonia | N | N | N |
| F | >65 to ≤70 | 6 | Covid-pneumonia | N | N | N |
| G | >35 to ≤40 | 9* | Covid-pneumonia | N | N | N |
| H | >45 to ≤50 | 4* | Covid-pneumonia | Y | N | N |
| I | >80 to ≤85 | 7 | Acute respiratory failure | N | N | N |
| J | >65 to ≤70 | 12 | Respiratory distress | Y | Y | N |
| K | >65 to ≤70 | 5 | Pneumonia | Y | N | Y |
| L | >60 to ≤65 | 6 | Dehydration | N | N | N |
| M | >50 to ≤55 | 5 | Covid-pneumonia | N | N | N |
| N | >60 to ≤65 | 7 | Covid-pneumonia | N | N | N |
| O | >55 to ≤60 | 6 | Pneumonia | N | N | N |
| P | >60 to ≤65 | 12 | Pneumonia | N | N | N |
| Q | >65 to ≤70 | 7 | Covid-pneumonia | N | N | N |
| R | >50 to ≤55 | 7 | Pulmonary embolism | N | N | N |
| S | >55 to ≤60 | 10 | Covid-pneumonia | N | N | N |
| T | >70 to ≤75 | 10 | Covid-pneumonia | Y | Y | Y |
| U | >35 to ≤40 | 6* | Covid-pneumonia | N | N | N |
| V | >80 to ≤85 | 8 | Dyspnea | N | N | N |
| W | >55 to ≤60 | 2 | Covid-pneumonia | Y | N | N |
| X | >50 to ≤55 | 7 | Covid-pneumonia | N | N | N |

ITT denotes intent-to-treat, N no, Y yes.

*The adverse event associated with the hospitalization started the day before the patient was admitted to the hospital.

**Figure S1. Patient disposition (ITT population).**

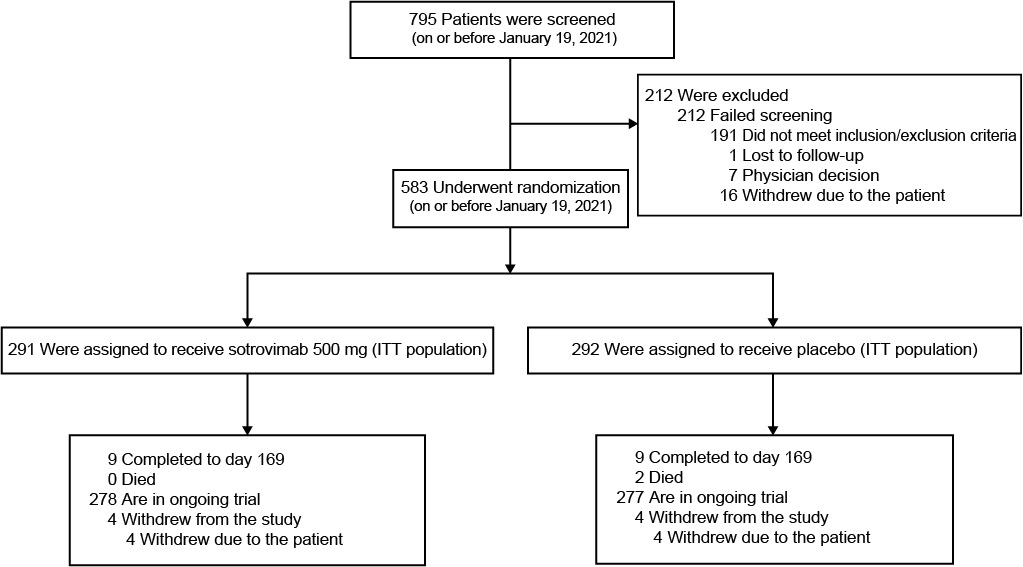

ITT denotes intent-to-treat.
